# The Ethnic/Racial Variations of Intracerebral Hemorrhage Genetics (ERICH-GENE) Study Protocol

**DOI:** 10.1101/2025.06.11.25329301

**Authors:** Guido J. Falcone, Stacey Q. Wolfe, Marialuisa Zedde, Rosario Pascarella, Jordi Jimenez-Conde, Marta Vallverdu Prats, Joan Jimenez-Balado, Alessandro Pezzini, Sandra Rossi, Rustam Al-Shahi Salman, Neshika Samarasekera, Ramin Zand, Jinag Li, Christina Jern, Daniel Strbian, Liisa Tomppo, Hanne Sallinen, Mar Hernandez Guillamon, Magdy Selim, Mayowa Owolabi, Rufus Akinyemi, Gregory Fakunle, Tsong-Hai Lee, David Werring, Isabel C. Hostettler, Henry Houlden, Pankaj Sharma, Isaac John, Gie Ken-Dror, Wendy Jenkins, Kevin N. Sheth, Lauren H. Sansing, Dharambir K. Sanghera, Evgeny Sidorov, Israel Fernandez-Cadenas, Jara Cárcel-Márquez, Ching-Jen Chen, Andrea Becerril-Gaitan, Keon-Joo Lee, Hee-Joon Bae, Martin Dichgans, Rainer Malik, Stephanie Debette, Aniket Mishra, Guillaume Pare, Michael Chong, Yoichiro Kamatani, Zhengming Chen, Robin G. Walters, Sudha Seshadri, Myriam Fornage, Catherine Sudlow, Lee A. Gilkerson, Vivek J. Khandwala, Thomas C. Maloney, Stacie Demel, Livia Parodi, Ramin Zand, Paul Nyquist, Wendy Ziai, Bradford Worrall, Vagal M Achala, Carl D. Langefeld, Jonathan Rosand, Christopher D. Anderson, Daniel Woo, the ERICH-Gene Investigators

**Affiliations:** Department of Neurology, Yale School of Medicine, New Haven, CT, USA; Yale Center for Brain and Mind Health, Yale School of Medicine, New Haven, CT, USA; Department of Neurosurgery, Wake Forest School of Medicine, Winston-Salem, NC, USA; Neurology Unit, Stroke Unit, Azienda Unita Sanitaria Locale IRCCS di Reggio Emilia, Reggio Emilia, Italy; Department of Neurology, IMIM-Hospital del Mar; Neurovascular Research Group, Institut Hospital del Mar d’Investigacions Mèdiques (IMIM), Barcelona, Spain; Department of Medicine and Surgery, University of Parma and Stroke Care Program, Department of Emergency, Parma University Hospital, Parma, Italy; Centre for Clinical Brain Sciences, University of Edinburgh, Edinburgh, UK; Department of Neurology, College of Medicine, The Pennsylvania State University, Hershey, PA, USA; Department of Neurology, Sahlgrenska University Hospital, Region Västra Götaland, Gothenburg, Sweden; Department of Neurology, Helsinki University Hospital and University of Helsinki, Helsinki, Finland; Neurovascular Research Laboratory, Vall d’Hebron Research Institute, Universitat Autònoma de Barcelona, Barcelona, Spain; Department of Neurology, Beth Israel Deaconess Medical Center/Harvard Medical School, Boston, MA, USA; Department of Medicine, University College Hospital, Ibadan, Nigeria; Stroke Center and Department of Neurology, Linkou Chang Gung Memorial Hospital, Taoyuan, Taiwan; UCL Queen Square Institute of Neurology, London, UK; Institute of Cardiovascular Research, Royal Holloway University of London, Egham, TW20 0EX, UK; Department of Pediatrics, College of Medicine, University of Oklahoma Health Sciences Center, Oklahoma City, OK, USA; Stroke Pharmacogenomics and genetics, Department of Neurology, Institut d’Investigació Biomedica Sant Pau, IIB-Sant Pau, Barcelona, Spain; Department of Neurosurgery, University of Texas Health Science Center, Houston, TX, USA; Department of Neurology, Korea University Guro Hospital, Seoul, Republic of Korea; Institute for Stroke and Dementia Research (ISD), University Hospital, LMU Munich, Germany; University of Bordeaux, INSERM, Bordeaux Population Health research center, UMR1219, Bordeaux, France; Population Health Research Institute, Hamilton Health Sciences and McMaster University, Hamilton, ON, Canada; Laboratory of Complex Trait Genomics, Department of Computational Biology and Medical Sciences, Graduate School of Frontier Sciences, University of Tokyo, Tokyo, Japan; Clinical Trial Service Unit, Nuffield Department of Population Health, University of Oxford, Oxford, UK; Glenn Biggs Institute for Alzheimer’s & Neurodegenerative Diseases, University of Texas Health Science Center at San Antonio, San Antonio, Texas, USA; Brown Foundation Institute of Molecular Medicine, McGovern Medical School, The University of Texas Health Science Center at Houston, Houston, Texas, USA; Centre for Medical Informatics, Usher Institute of Population Health Sciences and Informatics, University of Edinburgh, United Kingdom; Department of Neurology and Rehabilitation Medicine, University of Cincinnati College of Medicine, Cincinnati, OH, USA; Department of Radiology, University of Cincinnati College of Medicine, Cincinnati, OH, USA; Department of Neurology, Massachusetts General Hospital, Boston, MA, USA; Department of Neurology, Penn State College of Medicine, Hershey, PA, USA; Department of Neurology, Johns Hopkins School of Medicine, Baltimore, MA, USA; Department of Neurology, University of Virginia School of Medicine, Charlottesville, VA, USA; Department of Biostatistics and Data Science, Wake Forest University School of Medicine, Winston-Salem, NC, USA; Henry and Allison McCance Center for Brain Health, Massachusetts General Hospital, Boston, MA, USA; Department of Neurology, Mass General Brigham, Boston, MA, USA; Department of Neurology, Jacobs School of Medicine and Biomedical Sciences, University at Buffalo, Buffalo, NY

**Author notes:** **Correspondence: Christopher D. Anderson MD, MMSc**, 60 Fenwood Road, Boston, MA 02115, **Daniel Woo MD, MSc**, 1001 Main Street, Buffalo, New York 14203. Co-leading authors (Guido J. Falcone, Stacey Q. Wolfe). Co-senior authors (Christopher D. Anderson, Daniel Woo).

**Keywords:** Intracerebral hemorrhage, Hemorrhagic stroke, Population genetics, Genetics, Genomics, Genome-wide association study, Risk factors, Neuroimaging

## Abstract

**Background:** Spontaneous, non-traumatic intracranial hemorrhage (ICH) is highly heritable disease. However, the identification of the genetic risk factors driving this high genetic predisposition has been limited by small sample sizes and underrepresentation of non-European populations. The ERICH-GENE study will gather and harmonize clinical, neuroimaging and genomic data on the largest and more diverse collection of ICH cases assembled to date.

**Methods:** ERICH-GENE is an NIH-funded, multi-center, international, genetic and neuroimaging study that aims to achieve the necessary sample size and diversity required to accurately describe the genetic architecture and trans-ethnic variation of ICH. ERICH-GENE will collect and harmonize clinical, neuroimaging and genomic data at least 10,000 multi-ethnic ICH cases. These data will be aggregated with 20,000 existing ICH cases and 600,000 ICH-free controls available through completed studies by the International Stroke Genetics Consortium. To ensure validity, data will undergo extensive harmonization, including expert review of neuroimages to ensure spontaneous etiology and hemorrhage location. We will conduct genome-wide association studies of risk, severity and outcome of ICH, testing for effect modification by race/ethnicity, sex and hemorrhage location. We will also conduct pathway, polygenic risk score and Mendelian randomization analyses.

**Results:** This study will include whole genome sequencing data from 10,850 spontaneous ICH samples, including clinical and radiographic phenotypic data to ensure reliability of true non-traumatic, non-lesional ICH and lobar vs nonlobar location. Of these, 1,497 have already been genotyped using genome-wide arrays, 3,753 have undergone whole genome sequencing, and 5,600 will undergo genome-wide genotyping through ERICH-GENE. There are currently 42 contributing sites exceeding study milestone enrollments. 16,175 radiographic studies from 4,974 patients have been uploaded for harmonization to date, including 26% lobar and 64% nonlobar hemorrhages. Neuroimaging assessment will also include grading for white matter hyperintensities, cerebral atrophy, and presence and severity of IVH. Nearly 6,000 ICH cases will complete genotyping by August 2025. Data/material transfer agreements for summary statistics as well as additional samples are on target to meet the study’s objectives.

**Conclusion:** ERICH-GENE is the largest trans-ethnic genetic study of ICH conducted to date. Combining a diverse patient population with expert adjudication of neuroimaging data, ERICH-GENE will identify genetic risk loci that drive the high heritability observed for this disease and make a significant contribution to the understanding of the trans-ethnic variation of its genetic architecture.

## INTRODUCTION

Spontaneous, non-traumatic intracerebral hemorrhage (ICH) has the highest morbidity and mortality rate of any stroke subtype with about 79,000 new cases in the United States per year.^1^ About a third of ICH patients die during the hospital admission and another third remain severely disabled.^2^ There are clear disparities in ICH incidence, with higher risk of first-ever and recurrent ICH observed in Blacks, Hispanics and Asians. ^3–5^ The societal burden of ICH is likely to rise in coming years, based off clear trends over the previous two decades as well as increasing age and use of anticoagulant treatment.^1–3,6^ Unlike ischemic stroke, ICH has not shown any decline in observed incidence rate.^7–9^ However, ICH shares clinical, histopathologic and radiographic features within the spectrum of cerebral small vessel disease (CSVD), which contributes to stroke, depression, and Alzheimer’s Disease and related dementias.^10–14^

We now know that ICH is a complex disease with an estimated heritable component of 30%.^15^ Importantly, genetic variation appears to influence not only risk but also the acute severity and long-term clinical outcomes of ICH. We have previously identified novel genetic risk loci for ICH that have proven crucial to identifying underlying biological links between ICH, small vessel stroke, and CSVD. Our prior genome-wide association studies (GWAS) of ICH, some of which have combined ICH with related CSVD phenotypes, have found associations at *APOE*, *PMF1*, *ICA1L*, and *COL4A2*, providing insights into the roles of endothelial cell proliferation, basement membrane maintenance, and beta-amyloid in ICH risk.^14,16–18^ While initial discoveries were from predominantly White/Caucasian populations, subsequent analyses indicate substantial variability for associations across ethnicities.^16,19^ Black, Asian, and Hispanic populations suffer from ICH approximately 10 years earlier in life than whites, with disproportionate burden and effects of risk factors such as hypertension.^19^ Maximizing sample size as well as ethnic diversity when studying ICH genetics will enable the identification of new loci both within and between ethnicities, elucidating causal mechanisms and providing tools for genetic risk stratification in diverse patient populations.

As an example of the additional scientific value provided by studies with appropriate trans-ethnic representation, a recent meta-analysis confirmed the previously reported effect of *APOE* ε2 and *APOE* ε4 on ICH risk.^16^ However, when restricting the analysis to only self-identified Hispanics and blacks, attenuated effects were found, despite adequate power to detect an effect lower than that seen in whites.^16^ These results illustrate the variability of genetic effects in ICH by race and ethnicity and demonstrate the need for multi-ethnic samples to better understand how genetic risk interacts with other exposures. In this setting, the scientific goals of Ethnic/Racial Variations of ICH (ERICH)-GENE are to: 1) harmonize ICH case and exposure phenotypes, and perform a GWAS of ICH risk, CSVD imaging features, and outcome in a multi-ethnic dataset; 2) identify genetic risk factors for ICH that vary by sex or ethnicity, 3) build and validate genetic risk prediction tools for ICH risk and outcome, and 4) develop a data sharing platform incorporating whole genome sequencing and neuroimaging data.

## METHODS

Following recent guidelines of research terminology describing population descriptors,^20^ we will refrain from usage of the term “race”, which was originally included in the title of the ERICH-GENE study. To facilitate understanding, we will refer to “ethnicity” as a sociopolitical approximation of genetic ancestry, to which we here refer to as “ancestry”.

### The ERICH-GENE Study Group and objectives

The ERICH-GENE Study Group leverages existing and new collaborations to maximize efficiency and diversity in patient representation. ERICH-GENE combines ICH cases and controls that have already been genotyped with ICH cases and controls that will be newly genotyped. One batch of already genotyped participants is contributed by the ERICH study, that enrolled 3,000 ICH cases (1000 white, 1000 black and 1000 Hispanic) and 3000 matched controls at 41 sites across the US whose samples completed whole genome sequencing (WGS) as part of the NHGRI-sponsored Centers for Common Disease Genomics (CCDG) initiative^21^ A second batch of already genotyped cases is contributed by the Women’s Health Initiative (WHI), including an additional 753 ICH patients whose samples completed whole genome sequencing through the NHLBI-supported TOPMed initiative. In addition, 5,600 ICH cases are being newly enrolled and genotyped as part of ERICH-GENE. Importantly, these cases undergo expert review, harmonization and blinded case adjudication to assign hemorrhage location and exclude secondary causes of bleeding (e.g., tumors and vascular malformations). This process will allow us to better capture the two major pathophysiologic ICH mechanisms (hypertension-related vasculopathy and cerebral amyloid angiopathy) that underlie ICH and usually correspond to ICH location (nonlobar versus lobar, respectively). Finally, over 20,000 spontaneous ICH cases and 600,000 controls will be aggregated through summary statistics from various biobanks, including the UK Biobank, China Kadoorie Biobank, and Biobank Japan, and additional previously-genotyped cases and controls available through the Cerebrovascular Disease Knowledge Portal in association with the International Stroke Genetics Consortium. Rather than pursuing underpowered and unharmonized GWAS, we are performing a coordinated approach to maximize power for discovery of new loci for ICH.

### Definition of ICH

The primary phenotype is spontaneous, non-traumatic ICH, meeting the following eligibility criteria: 1) age ≥18 years, 2) associated acute symptoms (abrupt onset of severe headache, altered level of consciousness and/or focal neurologic deficit), 3) neuroimaging confirmation (head CT or brain MRI), and 4) no evidence of arteriovenous malformation, aneurysm, hemorrhagic conversion of ischemic stroke, trauma or brain tumor as a cause of the hemorrhage. ICH location, demographic and medical covariates, will be cataloged and harmonized by the Phenotyping Committee. On intake, phenotype libraries will be recoded using Common Data Elements to prevent phenotype misclassification and account for missing or discordant data elements between studies.

### Controls

Control subjects without ICH will be ethnically matched and randomly selected from the populations that gave rise to the cases. Both publicly available cohorts and ICH-free controls will be utilized from publicly available cohorts as well as from certain sites providing both ICH and ICH-free controls identified independently of the exposure. In total, the expected combined sample size of our analysis is 21,375 cases and 616,951 ICH-free controls, of which 8,953 cases and 30,971 controls from ERICH, WHI and ISGC will be included in the primary analysis stratified by location (lobar vs. nonlobar ICH, see Table 2).

**Table 1:**
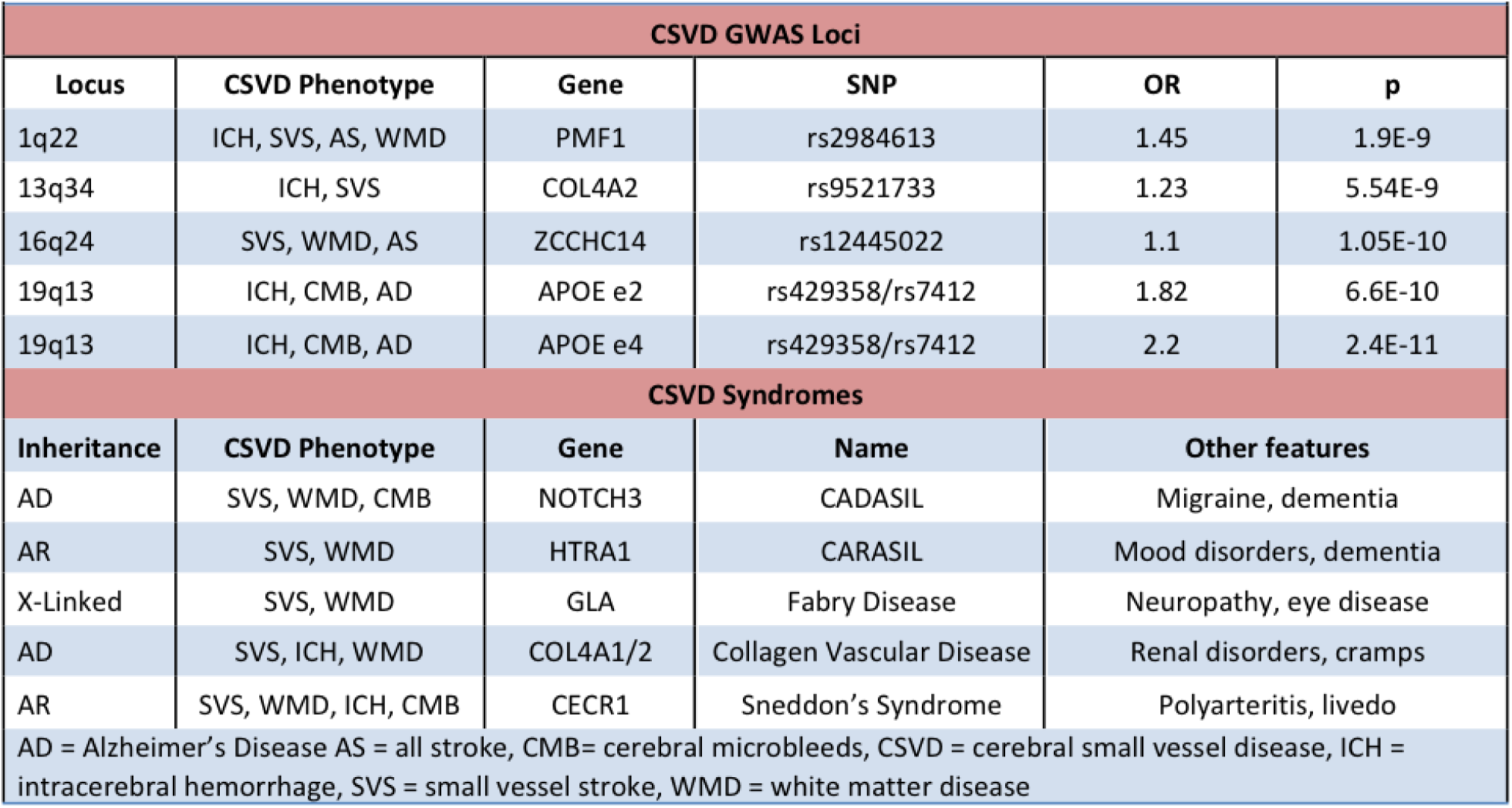
GWAS and Monogenic loci in ICH and CSVD.

**Table 2:**
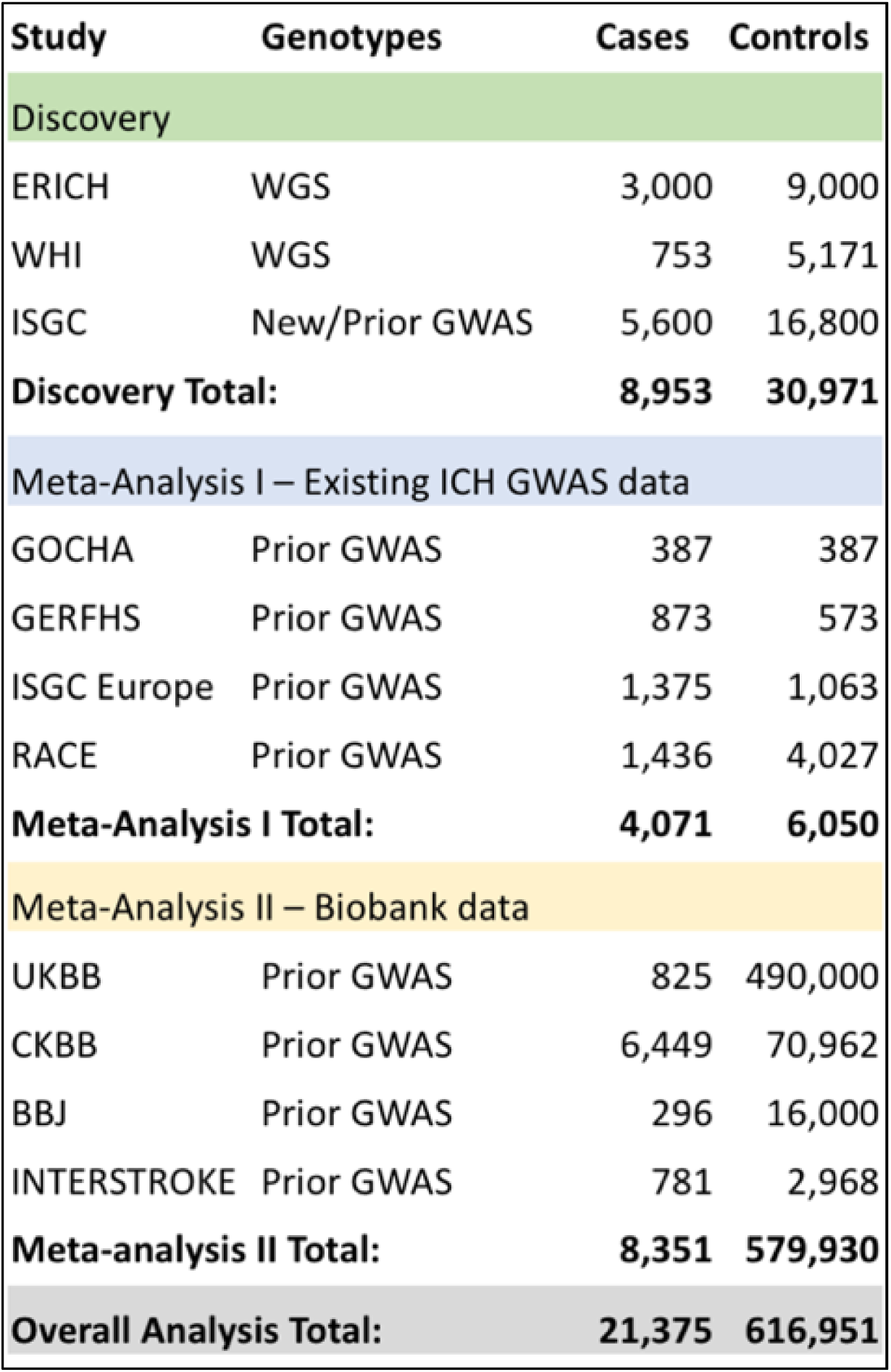
Overview of cohorts and sample sizes for ICH cases and controls. This table shows the sample sizes of cases and controls present or expected from each of the participating studies and their subdivision into discovery cohort, meta-analysis I and II.

### Neuroimaging and Imaging Validation

For validation of ICH as the qualifying event and reduction of heterogeneity in the ICH case population due to contamination with secondary causes of ICH, all collected neuroimaging will undergo central harmonization. All imaging will be submitted through Ambra (Intelerad, Montreal, Canada), a secure deidentified web-based portal, and reviewed by a panel of experts in ICH neuroimaging that includes stroke neurologists and neuroradiologist to confirm location and etiology. The location of ICH (nonlobar vs. lobar) is a crucial phenotype to ascertain, as hemorrhage location contains information about the underlying pathophysiology (i.e. hypertensive lipohyalinotic disease in the setting of nonlobar ICH, and cerebral amyloid deposition in the setting of lobar ICH) as well as the overall genetic architecture of the ICH trait. Additional CT-based measurements to be obtained include of ICH volume, presence and volume of intraventricular hemorrhage using the Graeb score, leukoaraiosis using the van Swieten scale, cerebral atrophy using the Global Atrophy Score. Where possible, we will adhere to NINDS Common Data Elements for collection of neuroimaging phenotypes. MRI will be solicited and available on a subset of patients for central collection and storage for confirmatory phenotyping and future initiatives.

### Clinical Data Acquisition

ERICH-GENE will collect core and secondary non-genetic variables. The group of core variables, were selected to facilitate the collection and harmonization of data across a large number of sites, include age, date of hemorrhage, sex, ancestry, ethnicity and whether admission CT is available. Secondary non-genetic variables will be collected when able, though not crucial for GWAS analyses, and include vascular risk factors (hypertension, diabetes, hypercholesterolemia, obesity, smoking), comorbidities (previous ICH or stroke and dementia), medications (antithrombotics and statins), and clinical variables and outcomes (admission NIHSS, Glasgow Coma Scale, and baseline, discharge, and follow-up mRS with emphasis at 3 months are collected).

#### Selection of Genotypes of Analysis

Genotypes for ICH cases will be ascertained using common variant data extracted from WGS in CCDG and TOPMed as well as a dedicated common variant GWAS in newly-genotyped individuals, with pre- and post-imputation harmonization. For all individuals with WGS-based genotyping, only common variants with minor allele frequency > 1% will be included in GWAS analyses. New GWAS genotyping in ERICH-GENE will be performed through the genotyping core at MGH with the Infinium Global Screening Array (GSA, Illumina, San Diego CA USA), performed at the Broad Institute. The GSA has 642,824 markers and was designed to accommodate multi-ethnic samples. At the conclusion of the study, all genotyped cases will be shared through the NIH-supported Genotypes and Phenotypes (dbGaP) platform as well as the ISGC-supported Cerebrovascular Disease Knowledge Portal (CDKP, https://cd.hugeamp.org/), in order to maximize the scientific impact across the research community.

### Statistical Methods

#### Quality Control Pipeline

Common variant QC approaches have become largely standardized and will be implemented pre- and post-imputation,^22–24^ including screening for batch effects and sex-inconsistency, and filtering for individual and variant missingness by study, differential missingness by case/control status, heterozygosity, Hardy-Weinberg Equilibrium, and identity by descent. Aligning strands across cohorts is critical and special attention will be given to single nucleotide polymorphisms (SNPs) that are A/T and C/G with minor allele frequency near 0.5. Population structure will be assessed at this step, using Eigenstrat, ADMIXTURE, and PC-AiR, and genetic outliers and those inconsistent with self-reported ethnicity will be removed.^25,26^

#### Variant imputation and harmonization

Using WGS and GWAS genotyping data, we will perform imputation to the 1000 genomes reference panel using IMPUTE2 within each dataset. Variants with high imputation quality (INFO > 0.5; confidence > 0.9) will be retained for analysis, resulting in a harmonized pool of common variants across included studies for uniform and unconfounded association testing. Common variants identified via WGS in ERICH and WHI but not included in the final imputed dataset from the new ISGC GWAS dataset will be excluded from further analysis. Downstream analyses will account for imputation uncertainty.

#### Single variant analysis

We will use logistic regression as well as logistic mixed modeling for single variant analyses using SNPTEST, PLINK and GMMAT in R,^27^ adjusting for covariates and accounting for imputation uncertainty. GMMAT incorporates population structure and a relatedness matrix, reducing false positive associations due to between-sample heterogeneity and ethnicity.^12^ Analyses will be performed within each self-reported ethnicity and within study, followed by meta-analysis via METAL.^28^ Benjamini-Hochberg false discovery rate adjusted p-values will be computed.^29^ Chromosome X variants will be tested.

#### Meta-analysis

We will employ a two-stage meta-analysis to account for heterogeneity in multi-ethnic results from primary association testing. We will cluster population subgroups with similar ancestral backgrounds prior to meta-analysis based on admixture estimates referent to anchoring populations (rather than a traditional cohort-based approach), followed by fixed-effects modeling using METAL.^28^ This approach has been shown to maximize statistical power while controlling Type I error due to heterogeneity in ethnically stratified datsets.^31^

#### Gene-based testing

Aggregation of variants by gene or other functional unit carries the advantages of limiting the number of statistical tests as well as combining the information from neighboring variants in a single gene into a variable with greater power. We will include both collapsing methods and sequence kernel association tests.^30^ Additional gene-set tools will also be employed.^31,32^ As with single variants, meta-analysis will be performed across ethnic groups and between studies, with multiple testing burden corrected using false discovery rate-adjusted p-values.

#### Outcome phenotypes

Prior ICH GWAS have successfully identified genetic loci associated with outcome,^33^ and modified Rankin Scale (mRS) and mortality at 3 months will be available for approximately 6,654 ICH cases in the present proposal. 3-month mRS will be binarized into good outcome (mRS 0-3) and poor outcome (4-6) in available participants. 1-year and further analyses of mRS will be converted to averaged z-scores to reduce bias due to floor/ceiling artifacts and tested using proportional hazards controlled for age, sex, and education. The ongoing Recurrent Hemorrhagic Stroke in Minority Populations study program is collecting long-term functional, cognitive, and psychiatric outcome data on 937 ERICH participants with > 4 years of average follow-up which will become available for genetic association testing during ERICH-GENE.

#### Genetic associations with neuroimaging phenotypes in ICH cases

Genetic variants will be tested for association with relevant neuroimaging phenotypes, including hemorrhage location l(lobar and nonlobar), hemorrhage volume, intraventricular hemorrhage, leukoaraiosis, and cerebral atrophy.

##### Polygenic risk scores (PRS) for ICH risk and outcomes

Using summary statistics from GWAS of ICH risk, we will extract ancestry-specific beta-coefficients, allele frequencies, and linkage disequilibrium (LD) information to construct PRS. This will maximize the predictive utility of the included variants based on regional associations rather than specific SNPs, which are more likely to vary across ethnicities.^34^ For each method, beta coefficients for all included variants are summed into a score by individual. Individuals with the greatest number of high-risk effect variants, in the tail of the distribution, represent the highest risk group for ICH risk or poor outcome compared with the rest of the population. This analysis will not only increase our understanding of the specific genetic locations that increase ICH risk across ethnicities, furthermore, it will provide a powerful clinical stratification tool (Figure 3).

#### Testing and validation of PRS performance

PRS for ICH risk and outcome will be validated in the meta-analysis and biobank datasets. Test characteristics for these PRS will be calculated in a minimal model, followed by inclusion of clinical and radiographic traits associated with ICH risk and outcome, to determine whether the PRS capture risk information orthogonal to classical risk factors. Risk and outcome PRS will also be applied within each representative ethnicity to determine utility across ancestries. Future analyses will include PRS generation within each ethnic strata combined with regional fine-mapping to optimize ICH PRS predictive utility for the broadest possible populations, a unique aspect to this application which addresses a key limitation in efforts to derive clinically meaningful PRS.

### Data sharing platform incorporating WGS and neuroimaging data

ERICH-GENE will support back-end development of WGS integration to update the CDKP, ensuring the NIH investment in this well-utilized resource remains modern and relevant.^35^ To date, the CDKP has been limited to GWAS statistics for case-control studies of stroke, as well as continuous phenotype GWAS of related traits such as BMI. We will integrate the ERICH genotypes and association results shared through the CDKP with CT-based phenotypes shared through the newly introduced, NIH-supported I-CDKP resource for neuroimage data sharing, in order to allow external investigators to access our adjudicated CT-based phenotypes, as well as to derive new image-based endophenotypes for further studies. Individuals in both portals will be given matching subject IDs to allow joint genetic-imaging analyses.

## STUDY UPDATE

ERICH-GENE received funding September 2021 and is currently in Year 4. A total of 42 institutions worldwide are enrolling in this study. Figure 1 provides an overview of the study timeline. In Years 1-3, the Ambra radiographic database has been up and running, and a total of 16,175 images from 4,974 patients with ∼ 3,000 MRIs are uploaded for expert review by the neuroimaging core, which is close to completion. In Year 4, an additional 4,000 cases will undergo phenotypic harmonization at the Cincinnati core. In Years 1-3, in addition to the 3,000 samples from ERICH and the 753 samples from WHI, ∼1,600 new ICH cases have received WGS, and an additional ∼2100 cases have been genotyped using the GSA. In Year 4, an additional 1,800 cases are pending genotyping from Asia, Europe and the US, followed by another estimated 2,000 cases from contributing sites in Africa, Europe, US and Asia.

**Figure 1.**
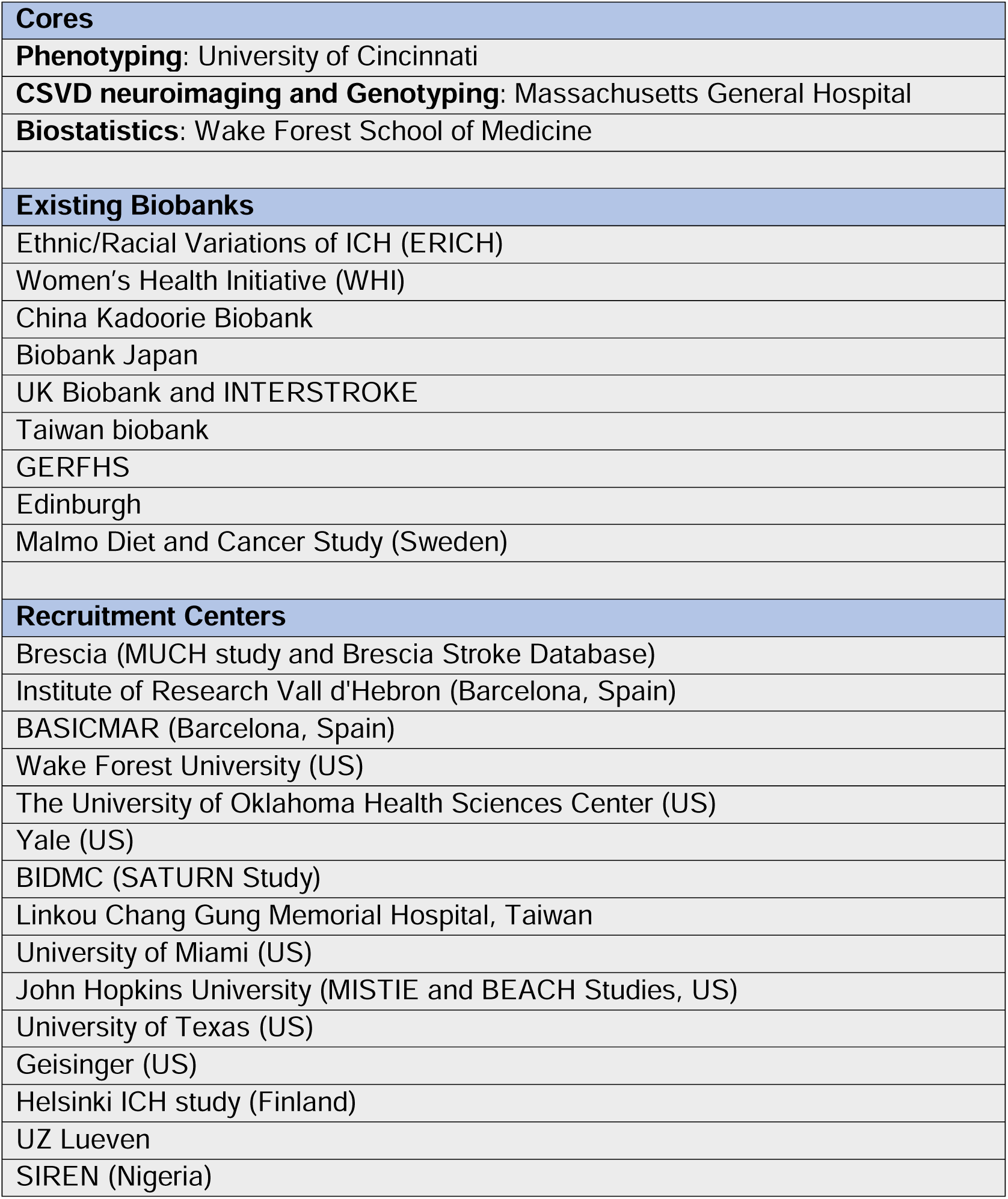
Study Organization and Contributors

**Figure 2.**
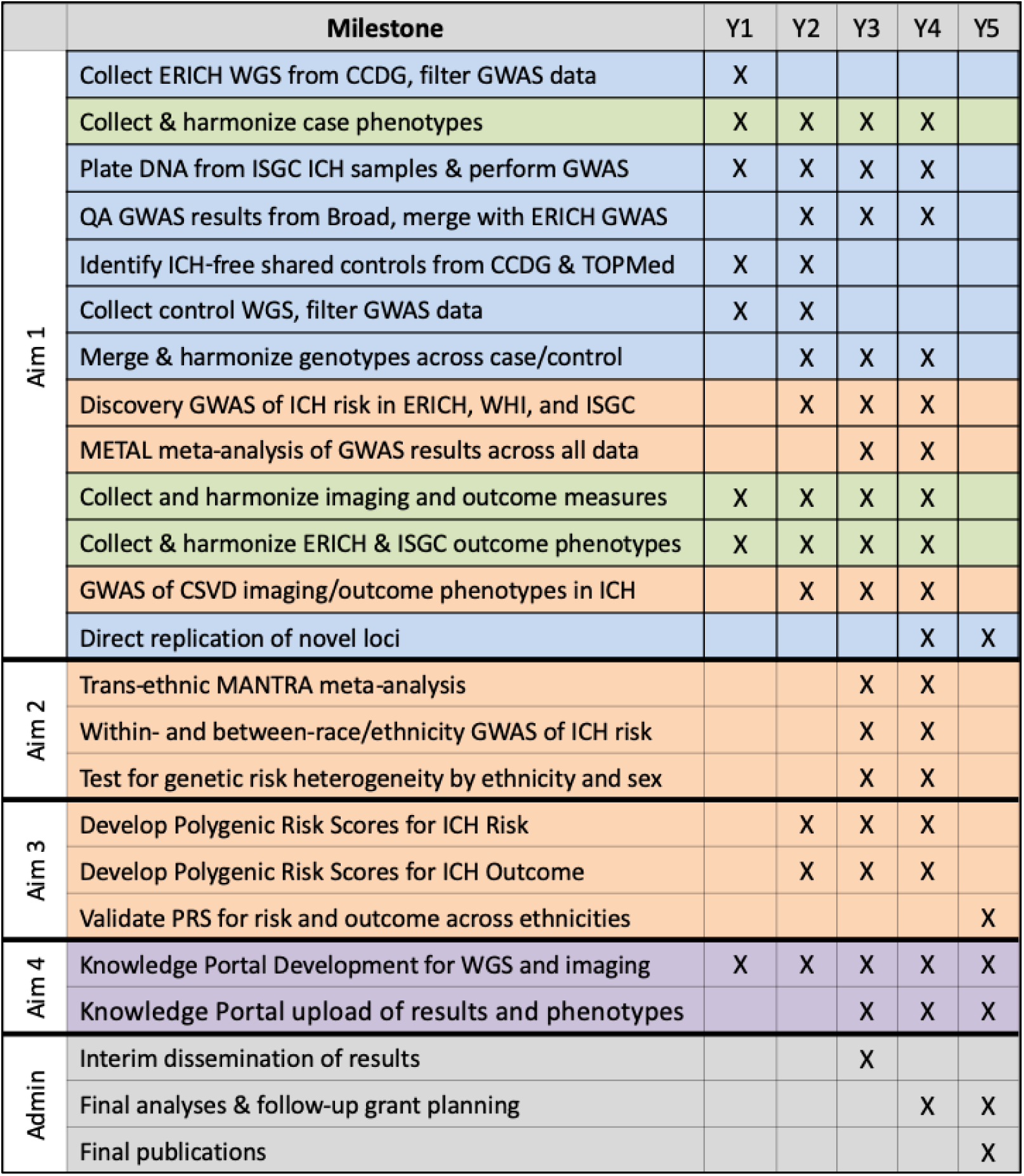
Study Timeline and Milestones

**Figure 3:**
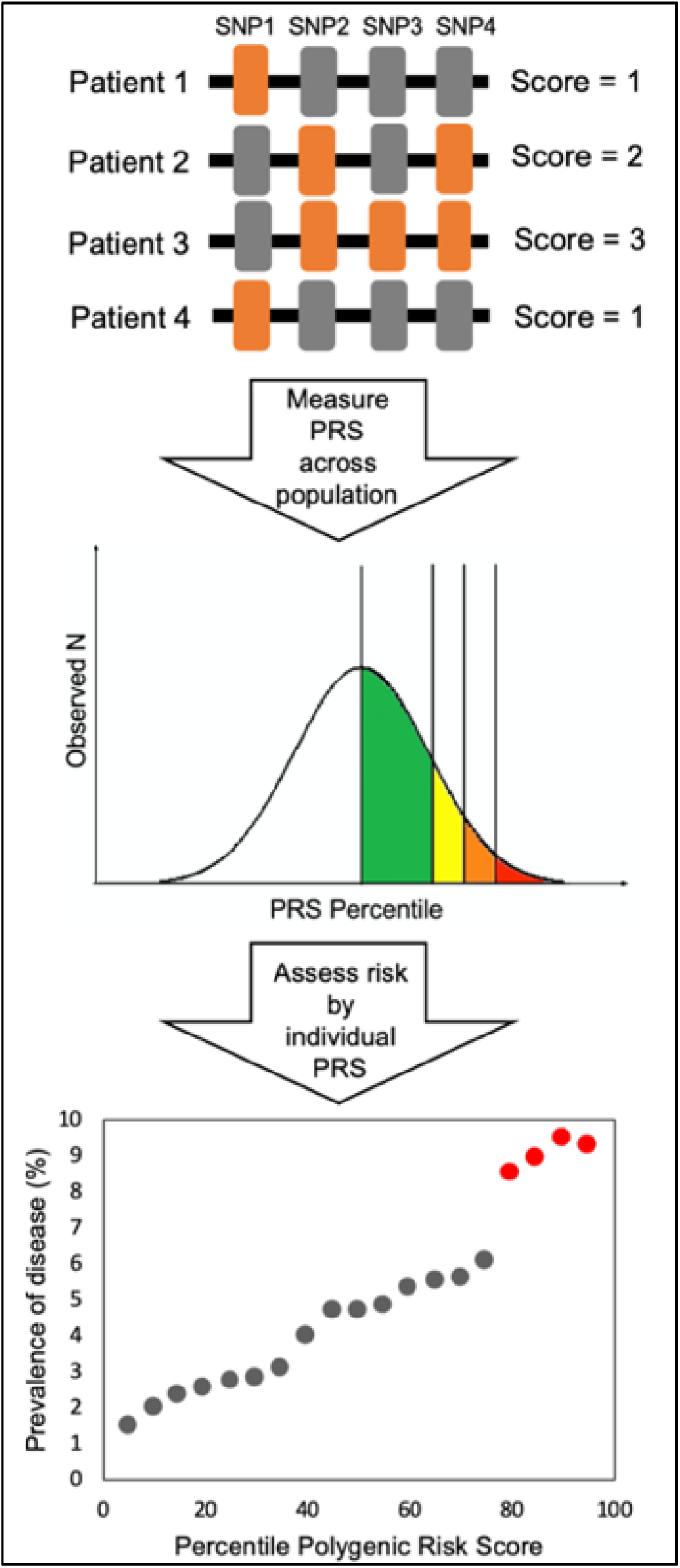
Design of Polygenic Risk Scores

## DISCUSSION

Despite current knowledge about the high heritability of ICH and the growing disease disparities across ancestries, research investigating the clinical and genetic predictors of ICH and outcomes remains limited by small study sizes and non-representative samples, primarily focused on white populations. In this study protocol, we introduce a collaboration of several of the largest studies and biobanks involving ICH patients, namely the ERICH study, WHI, ISCG contributors, China Kadoorie Biobank, Biobank Japan, and the UK Biobank. With the addition of 5,600 newly acquired ICH genotype and phenotype samples, all with centrally adjudicated neuroimaging, these combined data across many ethnicities will be used to perform the largest and most diverse harmonized GWAS to determine genetic risk in ICH to date, and to develop more specific and precise downstream genomic tools for clinical applications.

ICH is a devastating disease for patients, families and communities. It has the highest morbidity and mortality of all stroke subtypes, long hospital and rehabilitation length of stays, and significant loss of functional independence.^6^ There are remarkable differences in ICH risk across ethnic and socioeconomic strata. ICH incidence is 1.5-2-fold higher among African Americans and Hispanic Americans compared to White Americans.^21,36^ CSVD is highly linked to spontaneous ICH, with commonly found arteriolosclerosis (or lipohyalinosis) and cerebral CAA as underlying etiologies. Lipohyalinosis creates concentric hyalinized vascular wall thickening in the penetrating arterioles of the basal ganglia, thalamus, brainstem, and deep cerebellar nuclei with risk factors of hypertension, diabetes mellitus, and age. CAA is more often lobar and defined by arteriolar deposition of amyloid-β peptide. With approximately 30% heritability, associations at *APOE* (involved in the metabolism of lipids and implicated in both Alzheimer’s and cardiovascular disease), *PMF1* (involved in polyamine metabolism)*, ICA1L* (likely an autoantigen in diabetes and amyotrophic lateral sclerosis)*, and COL4A2* (found in collagen with the C-terminal portion an inhibitor of angiogenesis) have been found.^13–17,37^

Only recently has there been a renewed interest in aggressive treatment and study of this disease to impact patient outcomes. Surgical treatment in clinical trials such as MISTIE III and ENRICH have shown promising results,^38,39^ but treatments for prevention and secondary brain injury are clearly needed. Differences in outcomes amongst ethnicities, sexes, age and varying comorbidities must be addressed and understood. For example, mean age of ICH patients is lower among Black and Hispanic patients compared with White and over half of Black and Hispanic patients suffering ICH have untreated hypertension.^21^ The estimated yearly incidence of delayed dementia after ICH is 5.8%, which does not appear to correlate with hematoma location or size.^10^ Black and Hispanic ICH survivors had greater global small vessel disease on MRI as well as increased ICH recurrence.^40^ These data must also be considered in light of epigenetic factors such as 20-30% of underrepresented patients with ICH are uninsured, compared to 5% of White patients.^21^ The ERICH-GENE collaboration is an international commitment to analyze genomic risk factors which will impact and be key to both treatment development and outcomes. This effort will strive to overturn 25 years of failure for pharmacologic treatments in ICH and help identify and overcome treatment effect differences. Emphasizing the potential impact of genetic discoveries in terms of novel drug targets, recent findings indicate that pharmacological interventions with human genetic evidence supporting their mechanism of action have 2 times the chances of successfully moving from preclinical studies to FDA approval.

Genomic studies carry potential pitfalls that must be addressed. ERICH-GENE has developed both a genome and phenotype core, with centralized adjudicated neuroimaging to help ensure harmonization. Despite training of site-PIs enrolling in ERICH, centralized review ultimately excluded 6% of cases as not spontaneous ICH. Some of the most common misclassifications include hemorrhagic conversion of ischemic infarct, hemorrhagic metastases, contusion, ruptured aneurysm or AVM (Figure 4). In addition to case/not a case misclassification, ICH location determination errors were even more frequently identified with 211/3000 (9.3%) disagreement between the recruitment site and centralized reading and final adjudication. Because of the known differences in genetic architecture between lobar and nonlobar ICH, validation of neuroimaging is critical to minimize heterogeneity and enhance the potential to find true differences in a case-control study of validated spontaneous ICH.

**Figure 4.**
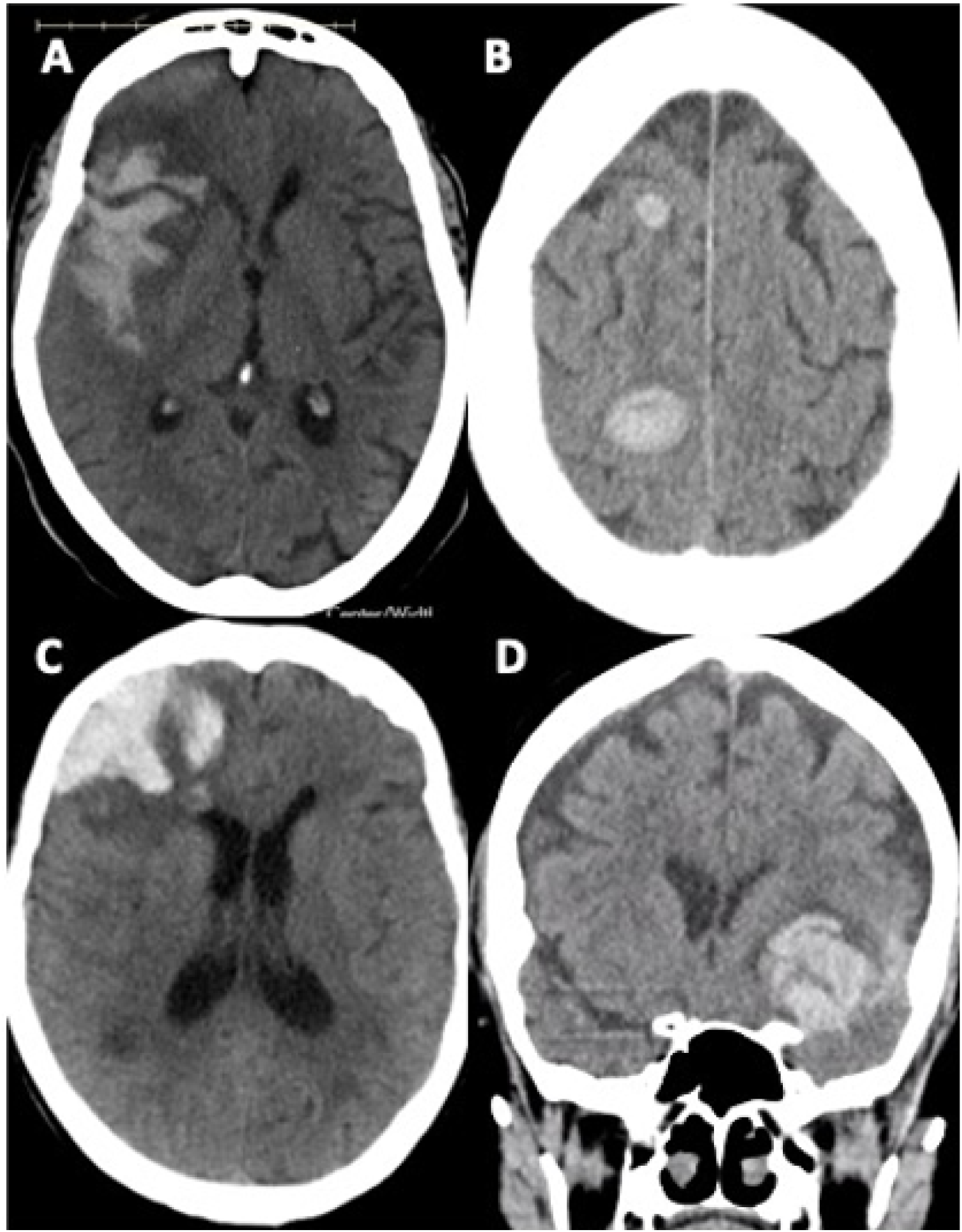
Examples of common ICH misclassifications. A) Submitted as lobar ICH, sharp delineation of hypodense borders extending to the cortex and less mass effect than expected for hemorrhage size indicate *hemorrhagic conversion of ischemic infarct*. B) Submitted as multiple amyloid ICH, however multiplicity and round shape of similar age bleeds indicate hemorrhagic metastases. C) Appearance consistent with contusion versus hemorrhagic infarct over spontaneous ICH. D) Submitted as deep ICH but location along the sylvian fissure is worrisome for a ruptured MCA aneurysm.

Novel genetic associations arising from ERICH-GENE carry great potential to refine the search for new therapeutic approaches, and to prioritize potential mechanisms for further study. Existing GWAS data in ICH has been crucial in identifying the pathobiological links between nonlobar ICH, small vessel ischemic stroke, and CSVD phenotypes such as white matter hyperintensities on MRI. These phenotypes have been epidemiologically linked for many years, but genetics offers a key opportunity to demonstrate shared biological origins. Furthermore, methods that build on GWAS data, such as Mendelian Randomization, have provided valuable new information about the causality of risk factors that association with ICH risk, including lipid levels, chronic kidney disease, statin exposure, and obesity. Finally, the construction of polygenic risk scores using existing ICH GWAS data have recently shown great promise in identifying individuals at the greatest risk for ICH, even beyond traditional clinical risk factors, and the association with ethnicity and epigenetics. For example, unlike White patients, *APOE* alleles were not associated with lobar ICH among Black or Hispanic individuals.^21^ Each of these efforts will be greatly strengthened through the influx of additional well-phenotyped ICH cases amassed under ERICH-GENE, which will drastically improve statistical power and permit new exploration of multi-ethnic models.

### Potential Limitations

Our nested case-control study design allows us to maximize our sample size, while accounting for differences in the population by the above-mentioned harmonization strategies. However, some ethnicity-specific effects of small effect size may escape detection. Our expected power would allow us to detect some variants with modest effect sizes, while our trans-ethnic meta-analysis will allow for maximal power exploring novel ICH associations.

The opposite, a lack of ethnicity-specific risk factors for ICH, might also be an obstacle. Given our preliminary data on *APOE*^16^ and the success of trans-ethnic studies of related neurologic diseases of aging such as Alzheimer’s Disease^14^ however, this is an unlikely risk. Furthermore, a lack of ethnicity-specific loci for ICH risk would provide new insight into the genetic architecture of ICH and provide broad support for an inclusive strategy for ICH case aggregation in future studies of ICH.

GWAS identify genetic associations with disease, but assumptions about causality cannot be made. Non-causal genetic factors are still valid in predicting and stratifying risk in populations through PRS, and growing availability of WGS data will allow exploration of risk loci via fine-mapping and functional modeling. Given that WGS is available for the ERICH participants, we will lay the groundwork to perform multi-ethnic fine-mapping to detect causal loci.

Protocols building on our results will be able to perform WGS or targeted sequencing on loci arising from our GWAS. The Multiplexed analysis of variant effects in cell and tissue models is already being leveraged in analyses of existing CSVD loci. Variants arising from our study will represent exciting new candidates for in vitro exploration using reporter assay and gene-editing studies.

Furthermore, future clinical practice will increasingly incorporate PRS into medical decision-making. Because of known degradation of predictive utility of PRS outside of ethnic populations in which they are discovered, it is critical that PRS for ICH be constructed using the most ancestrally diverse population possible. Well-characterized PRS for ICH risk and outcome will require validation in prospective populations and determination of best practices for PRS measurement, disclosure of results, and integration into health records. Our study will advance the usage of genetic information into clinical care in ICH, in concert with mitigation of other risk factors such as hypertension, sleep apnea, and socioeconomic factors,^21^ as well as concurrent efforts across other cardiometabolic diseases.^41,42^

## CONCLUSION

The ERICH-GENE study will provide the largest and ethnically most diverse dataset of genetic determinants of ICH. It will not only incorporate clinical and WGS data of ICH patients and controls, but also harmonized and adjudicated neuroimaging and clinical outcome data, that will be made openly accessible as an online data repository. Out study will address the increasing need of building diverse and comprehensive datasets as a basis for GWAS to provide information about genetic risk factors of ICH representative of a whole population and shed light on the genetic predictors of ICH and their clinical application.

## Data Availability

All data produced in the present study are available upon reasonable request to the authors.

## ACKNOWLEDGEMENTS

We would like to thank the patients and families that agreed to participate in this study. We would also like to thank all the research teams from the numerous institutions around the world that have made this study possible.

